# SARS-CoV-2 Infection and Stroke: Coincident or Causal?

**DOI:** 10.1101/2020.07.17.20156463

**Authors:** Melanie Walker, Christopher C. Young, Malveeka Sharma, Michael R. Levitt, David L. Tirschwell, the WWAMI Stroke Investigators

**Author notes:** **Corresponding Author**: Melanie Walker, MD, c/o Neurosurgery Publications, Department of Neurological Surgery, Harborview Medical Center, University of Washington, 325 9th Avenue, Box 359924, Seattle, WA 98104. **Disclosure statement**: None.

## Abstract

Neurological manifestations of SARS-CoV-2 infection described in isolated case reports and single institutions do not accurately reflect the clinical spectrum of disease across all geographies in a global pandemic. Data collected during peak of the Covid-19 pandemic from stroke centers in five states reveal few similarities to what has recently been published. Given the diversity in phenotype, we caution policymakers and health care providers when considering cerebrovascular complications from SARS-CoV-2 infection.

## Introduction

An associated hypercoagulable state has been implicated as the cause for stroke in as many as 5.9% of severely infected patients in Wuhan, China [1], 0.9% of patients hospitalized patients in a New York City hospital system [2], and as an unusually common presentation for young adults in New York City [3]. None of these findings were substantiated in a population of 11.8 million people across the five-state (Washington, Wyoming, Alaska, Montana, and Idaho [WWAMI] region in the United States.

## Methods

Nineteen of twenty-one nationally or state certified thrombectomy capable stroke centers in the WWAMI region provided data on stroke admissions and mechanical thrombectomies performed during March and April 2020 and March and April 2019 (Table 1). With the exception of Wyoming, all states were under shelter-in-place or stay-at-home mandate during this period of time.

**Table 1.** Stroke volume and treatment trends during Covid-19 pandemic in WWAMI thrombectomy centers. Values expressed as number of patients and standard deviation (SD). *One-sample t-test with statistical significance with >99% confidence level.

|  | Month |  |  |  |
| --- | --- | --- | --- | --- |
|  | March 2019 | April 2019 | March 2020 | April 2020 |
| Total ischemic strokes admissions | 956 | 859 | 828 | 630 |
| Average ischemic stroke admissions at each center (SD) | 53.1 (34.1) | 47.7 (32.6) | 46.0 (33.2)<br>p=0.53 | 37.1 (28.6)<br>p=0.31 |
| Average ischemic stroke admissions at each center expressed as a percentage of the monthly total in the preceding year (SD) | - | - | 90.7 (32.0)<br>p=0.23 | 81.3 (22.8) *<br>p=0.003 |
| Total number of mechanical thrombectomy procedures | 53 | 65 | 66 | 53 |
| Total SARS-CoV-2 patients undergoing mechanical thrombectomy | 0 | 0 | 1 | 0 |
| Average number of mechanical thrombectomy procedures at each center (SD) | 2.9 (2.6) | 3.6 (2.6) | 3.7 (3.6)<br>p=0.45 | 3.1 (2.2)<br>p=0.53 |

## Results

Compared to March and April 2019, we observed a 13.3% and 26.7% decrease in ischemic stroke admissions across the WWAMI region in March and April 2020, respectively. To control for disproportionate impact of changes at a few large centers, we analyzed average stroke volume at each stroke center as a percentage of the preceding year and found a statistically significant decrease in stroke volume in April 2020 compared to 2019 (18.7% decrease, p=0.003). The total and average number of mechanical thrombectomy procedures performed at each center during the pandemic were not significantly different compared to 2019. There was a trend towards higher average National Institute of Health Stroke Scale score at presentation (6.8 vs 7.2 in 2019 and 2020, respectively). Of 18,070 confirmed SARS-CoV-2 infections in the WWAMI region as of April 30, 2020, fifteen patients (<0.1%) were diagnosed with concomitant acute ischemic stroke; of these, one underwent mechanical thrombectomy and two were under the age of 50. There were no reports of venous sinus thrombosis in SARS-CoV-2 patients.

## Discussion

Consistent with global reports, we observed a regional reduction in overall stroke volume during the COVID-19 pandemic peak. Surprisingly, less than 0.1% of patients suffered coincident SARS-CoV-2 infection and ischemic stroke. We also found no association between SARS-CoV-2 infection and stroke in younger patients. Though the total number of ischemic stroke patients decreased, likely due to social distancing and changes in health-seeking behavior, the number of large vessel occlusions requiring thrombectomy was not significantly different. Our results suggest striking differences in the effects of COVID-19 on the presentation of ischemic stroke. The pathophysiological basis for these differences is unknown, and potentially related to differences in geography, viral strain, or other yet unidentified factor(s). Similarly, a retrospective review in the UK during this same period [4] failed to identify a causal relationship in six patients with large vessel strokes and coincident SARS-CoV-2 infection because competing vascular risk factors were present.

## Conclusion

Regional data from five U.S. states suggests that among patients who sought care or were hospitalized during the peak of COVID-19, acute ischemic stroke in the setting of SARS-CoV-2 infection is rare and may be coincident. Further study is necessary, and we caution clinicians making major clinical decisions based on isolated case reports involving single institutions.

## Data Availability

Data is found in the table in the manuscript and available in an excel spreadsheet if requested.

## WWAMI Stroke Investigators

<u>University of Washington Harborview Medical Center, Seattle, WA.</u> Melanie Walker, M.D., Christopher Young, M.D. Ph.D., Michael R. Levitt, M.D., Malveeka Sharma, M.D. M.P.H., Tricia O’Donohue, M.P.H., B.S.N., R.N., and David Tirschwell, M.D. M.Sc.

<u>Providence Alaska Medical Center, Anchorage, AK.</u> Judy M. Hayes D.N.P., A.P.R.N., A.C.N.S.-B.C., C.M.S.R.N.

<u>Alaska Regional Hospital, Anchorage, AK.</u> Sylvia Mau, R.N.

<u>Saint Alphonsus Regional Medical Center, Boise, ID.</u> Jane K. Spencer, A.C.N.S.-B.C., S.C.R.N., P.C.C.N.

<u>Eastern Idaho Regional Medical Center, Idaho Falls, ID.</u> Cheri Arnold R.N.

<u>St. Luke’s Boise Medical Center, Boise, ID.</u> Amy Morgan R.N.

<u>Evergreen Hospital Medical Center, Kirkland, WA.</u> Jennifer Griner, R.N., B.S.N., M.B.A. and David J. Likosky M.D.

<u>MultiCare Deaconess Hospital, Spokane, WA.</u> Jon E. Ween, M.D.

<u>MultiCare Tacoma General Hospital, Tacoma, WA.</u> Phyllis Smith, R.N.

<u>Overlake Hospital Medical Center, Bellevue, WA.</u> Jennifer Kurtz, M.S. C.C.C.-S.L.P.

<u>Providence St. Peter Hospital, Olympia, WA,</u> Tracey Ash, R.N., M.S.N., Joseph Ho, M.D. PhD., Madeline Nguyen, M.D.

<u>St. Joseph Medical Center (CHI Franciscan Health), Tacoma, WA.</u> David M. Lundgren and Anna Moore

<u>Southwest Medical Center (Peace Health), Vancouver, WA.</u> Kimberly Kirkpatrick, M.S., R.N.

<u>Swedish Cherry Hill, Seattle, WA.</u> Bronwyn S. Rogers B.S.N., R.N. and Yince Loh, M.D.

<u>University of Washington Valley Medical Center, Renton, WA,</u> Erin Eddington Alden, A.R.N.P.-C.N.S., A.C.N.S.-B.C.

<u>Virginia Mason Medical Center, Seattle, WA.</u> Fatima Milfred, M.D., Robert W. Ryan, M.D., M.Sc., F.R.C.S.C. and Allison Cotterman, R.N. B.S.N.

<u>Madigan Medical Center, Tacoma, WA.</u> Kari Heber, M.D.

<u>Wyoming Medical Center, Casper, WY.</u> Melody Bowar, C.N.R.N., S.C.R.N.

## References

1. Mao L, Jin H, Wang M, et al. Neurologic Manifestations of Hospitalized Patients with Coronavirus Disease 2019 in Wuhan, China [published online ahead of print, 2020 Apr 10]. JAMA Neurol. 2020; e201127. doi:10.1001/jamaneurol.2020.1127.

2. Yaghi S, Ishida K, Torres J, et al. SARS2-CoV-2 and Stroke in a New York Healthcare System [published online ahead of print, 2020 May 20]. Stroke. 2020; STROKEAHA120030335. doi:10.1161/STROKEAHA.120.030335.

3. Oxley TJ, Mocco J, Majidi S, et al. Large-Vessel Stroke as a Presenting Feature of Covid-19 in the Young. N Engl J Med. 2020;382(20): e60. doi:10.1056/NEJMc2009787.

4. Beyrouti R, Adams ME, Benjamin L, et al. Characteristics of ischaemic stroke associated with COVID-19 [published online ahead of print, 2020 Apr 30]. J Neurol Neurosurg Psychiatry. 2020; jnnp-2020–323586. doi:10.1136/jnnp-2020-323586

